# Inactivated Japanese encephalitis virus vaccination imprints fusion loop–biased antibody responses that are attenuated by repeated live-attenuated dengue vaccination

**DOI:** 10.64898/2026.02.27.26347269

**Authors:** Patrick I. Mpingabo, Esther I. Adekomi, Lisa A. Ware, Monir Hossain, Joseph Q. Lu, Heather Friberg, Gregory D Gromowski, Kathryn B. Anderson, Stephen J. Thomas, Adam T. Waickman

## Abstract

Immune imprinting, also known as immune history, is a core aspect of adaptive immunity that influences antibody responses to future antigen exposures. Nevertheless, the impact of sequential flavivirus vaccinations on epitope targeting and antibody activity in humans remains incompletely understood. This question is particularly important in regions where the inactivated Japanese encephalitis virus (JEV) vaccines and the live-attenuated dengue virus (DENV) vaccines are used, as both have been associated with an increased risk of symptomatic dengue infection and severe illness. We studied the impact of prior inactivated JEV IXIARO vaccination and simultaneous vaccination on humoral immunity following live-attenuated dengue CYD-TDV vaccination. Long-term analysis showed that JEV IXIARO priming guides the dengue vaccine–induced antibody response toward conserved fusion loop epitopes (FLEs) of the DENV envelope protein, as indicated by 4G2 FLE-bias. This imprinting was characterized by higher levels of 4G2 FLE-like antibodies, rapid recall responses after dengue vaccination, and broad but low-potency neutralization across dengue serotypes and Zika virus. Notably, 4G2 FLE–focused responses correlated with higher FcγRIIa-mediated antibody-dependent enhancement relative to neutralization potency, suggesting functional effects beyond neutralization. To better understand epitope dominance within the native envelope, we used a structurally defined fusion loop epitope mutant (FLE-mut) envelope dimer assay. Disrupting fusion loop accessibility significantly decreased antibody binding, confirming that FLE-specific antibodies are a major component of the response after sequential vaccination. Importantly, a complete series of live-attenuated dengue vaccine reduced 4G2 FLE bias, encouraged the recruitment of non-fusion-loop epitopes, and lessened FcγRIIa-biased antibody activity. Overall, these results show that vaccination platform, timing, and regimen are critical determinants of epitope dominance and antibody quality following flavivirus vaccination.

## INTRODUCTION

Flaviviruses such as the dengue viruses (DENVs), Japanese encephalitis (JEV), West Nile (WNV), yellow fever (YFV), and tick-borne encephalitis viruses (TBEV) are important human pathogens responsible for severe systemic and encephalitic diseases^1,2^.

Antibody-mediated immunity plays a vital role in defending against viral infections and is a key target of vaccination strategies. Although vaccine-induced antibodies are primarily assessed by their capacity to neutralize viruses, they also possess various Fc receptor–dependent functions that impact pathogen interaction, viral entry into cells, and activation of immune responses. Increasing evidence indicates that the epitope specificity of an antibody, rather than its abundance, is crucial in shaping its function and influencing biological and clinical outcomes.

A key aspect of adaptive immunity is immune imprinting, also known as original antigenic sin (OAS), in which prior exposure to an antigen influences responses to related future antigens by activating memory B cells^3,4^. Numerous studies on flaviviruses, including JEV, DENV, and ZIKV, have examined how prior immune encounters affect antibody responses and disease severity. These investigations show how past infection or vaccination shapes responses to subsequent infections and their outcomes ^5–9^. Although immune imprinting has been widely studied across various viral infections and vaccines, including influenza^4,10^, HIV^11^, SARS-CoV-2^12–16^, and DENV^17,18^, the detailed mechanisms by which it affects antibody epitope specificity and function after flavivirus vaccination remain incompletely understood. This knowledge gap is particularly significant in regions where multiple flaviviruses co-circulate and sequential immunization with different vaccines, such as JEV and DENV, is common.

JEV and DENV share significant structural similarities in their envelope (E) glycoproteins, essential for viral attachment and membrane fusion. The fusion loop epitope (FLE) located in domain II (EDII) of the E protein is highly conserved across flaviviruses and serves as a key target for cross-reactive antibodies. Conversely, domain III (EDIII) of the E protein is targeted by type-specific (TS) antibodies, which confer durable neutralizing immunity against the same virus ^19–22^. Antibodies against the FLE tend to be broadly cross-reactive but only weakly neutralizing ^23^, and they participate in Fc receptor–dependent functions, including antibody-dependent enhancement (ADE) observed *in vitro* ^24–26^. Cross-reactive murine monoclonal antibodies, such as 4G2 and 4E11, are widely used as molecular probes to identify responses specific to the EDII fusion loop and EDIII, respectively, thereby aiding epitope mapping^27–29^. Most prior studies on FLE reactivity use peptide-based or denatured-antigen assays that do not distinguish epitope dominance within the native E protein structure. To address this, a structurally defined envelope dimer protein with a FLE mutation at position 106 (G106D) was created to selectively hinder FLE accessibility while preserving the overall E-dimer structure ^30,31^. The FLE mutant (FLE-mut) E-dimer construct (SC.10) enable direct evaluation of FLE dependence by specifically disrupting the antigen structure rather than relying on linear epitope binding ^32,33^.

Dengvaxia® is a live-attenuated, tetravalent dengue vaccine based on the yellow fever 17D backbone, known as CYD-TDV. It expresses the premembrane (prM) and envelope (E) proteins of all four DENV serotypes^34^. While CYD-TDV elicits neutralizing antibodies, its effectiveness and safety depend on the individual’s baseline dengue serostatus^35^, underscoring the role of preexisting immunity in vaccine outcomes. Conversely, IXIARO, a Japanese encephalitis virus vaccine, is an inactivated vaccine produced in Vero cells^36^ that induces strong immune responses, generating protective antibody levels following a primary vaccination series in over 99% of children and adults, with slight variations by age. Although immunity declines over time^37^, booster doses can elicit a robust memory response, extending the period of protection. Moreover, inactivated JEV IXIARO vaccine can induce cross-reactive antibodies, suggesting they primarily expand memory B cells targeting conserved epitopes ^38^. In contrast, CYD-TDV, a live-attenuated dengue vaccine, provides continuous antigen exposure through limited replication, possibly further broadening B cell responses. It remains to be seen whether prior vaccination with inactivated JEV IXIARO promotes the development of conserved FLE–biased antibody responses after CYD-TDV vaccination and how such imprinting might affect antibody functions, as this has not been systematically studied in humans.

In this study, we examined whether prior vaccination with inactivated JEV IXIARO affects the humoral immune response by promoting conserved 4G2 FLE-specific responses, which are more likely to be recalled after CYD-TDV vaccination. Using longitudinal samples and combined analyses of epitope specificity, neutralization capacity, and FcγRIIa-mediated ADE, we found that previous JEV IXIARO vaccination skews the antibody response toward the 4G2 FLE. This response is characterized by reduced binding when the FLE is structurally altered, broad yet low-potency neutralization, and increased FcγRIIa-mediated ADE activity. Notably, our results also suggest that higher antigenic stimulation from subsequent full, 3-dose series of live-attenuated dengue CYD-TDV vaccination can partially overcome this 4G2 FLE imprinting, promoting the development of non–fusion loop epitopes and reducing ADE mediated by 4G2 FLE bias. Overall, our findings indicate that 4G2 FLE imprinting results from sequential flavivirus vaccination, and factors such as vaccination timing, regimen, and vaccine platform can influence vaccine-induced antibody response.

## RESULTS

### Study design and participants

Adults aged 18 to 45 with no previous flavivirus exposure participated in a phase II, randomized, open-label study conducted at a single center to assess the live-attenuated Chimeric Yellow Fever-Derived Tetravalent Dengue vaccine (CYD-TDV, Dengvaxia). The trial compared two vaccination schedules: the standard (0, 6, and 12 months) and an accelerated (0, 2, and 6 months) schedule, with the vaccine administered alone, in combination, or sequentially with a purified inactivated JEV vaccine (IXIARO)^39^. For this study, sera from participants who received the accelerated CYD-TDV schedule (0, 2, and 6 months), either alone or combined with IXIARO, were selected for analysis. Samples were collected at baseline (day 0), as well as on days 28, 84, 196, and 336 after vaccination (**Fig. 1A**). Participants were categorized based on prior JEV IXIARO vaccination status: Group 1 included JEV-naïve individuals who received only CYD-TDV (**DV-CYD**); Group 2 included those who received the first CYD-TDV dose simultaneously with JEV IXIARO (**JE IXIARO+DV-CYD**); and Group 3 consisted of individuals who primed with JEV IXIARO 6 months prior receiving CYD-TDV (JE IXIARO-DV-CYD) (**Fig. 1A**). Longitudinal serum samples were tested for antibodies to DENV virus-like particles (VLPs), antibodies to YFV non-structural-1 (NS1) protein, JEV VLP-reactive IgG, DENV VLP-reactive IgM, broad flavivirus neutralization, and FcγRIIa-mediated antibody-dependent enhancement (ADE) activity.

**Figure 1.**
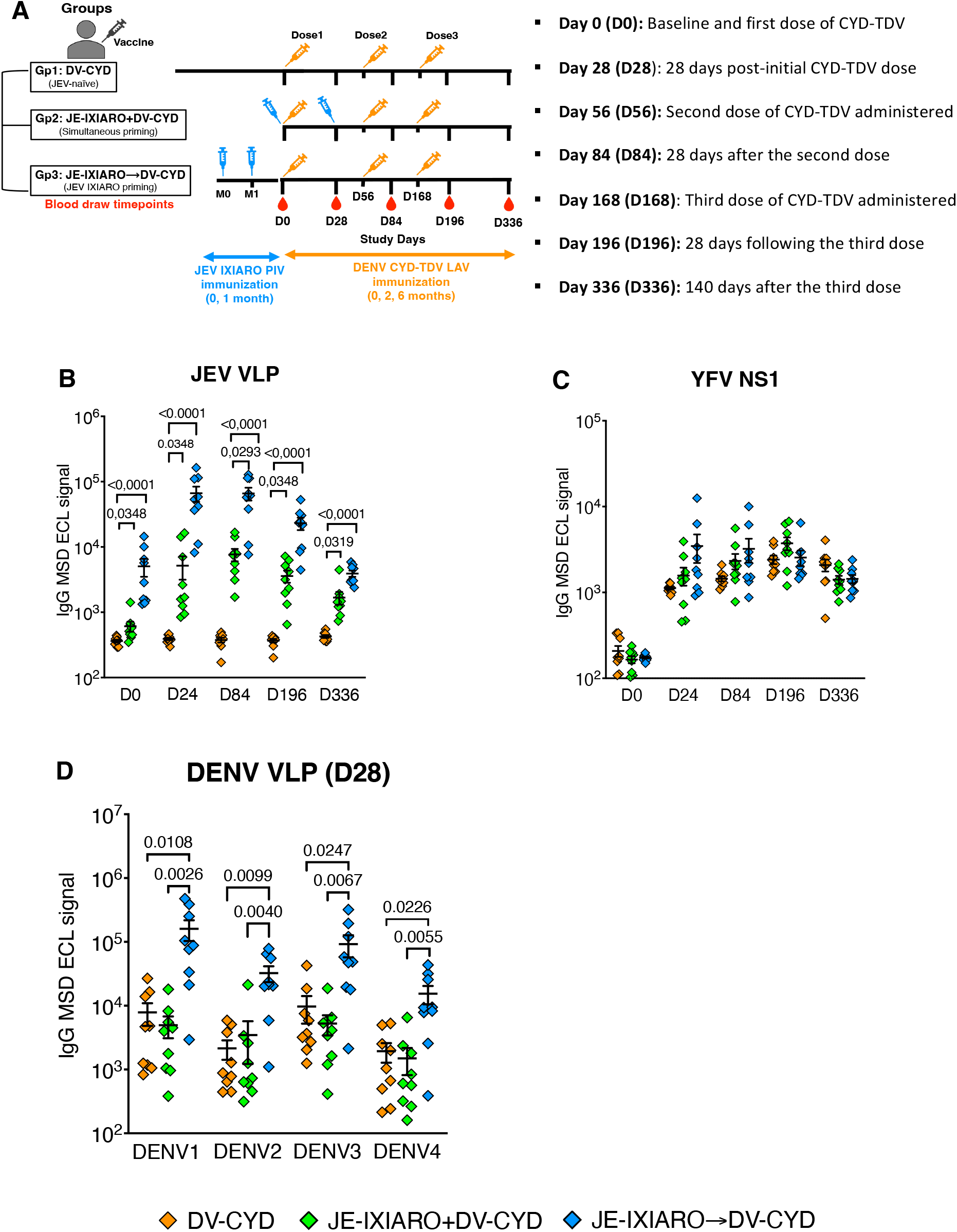
Study design, vaccination regimens, longitudinal sampling, and vaccine antigens-reactive IgG response. **(A)** An overview of the study design and vaccination strategies. Participants were categorized by their Japanese encephalitis virus (JEV) vaccination history as JEV-naïve (n=9), sequentially primed with inactivated JEV IXIARO followed by dengue CYD-TDV (n=9), or simultaneously administered both vaccines within the same immunization window (n=9). All participants received a full dose of live-attenuated dengue CYD-TDV vaccine. Blood samples were taken at baseline (day 0, before dengue vaccination) and on days 28, 84, 196, and 336 after vaccination. Serum samples underwent IgG-- and IgM-binding assays, epitope-specific blockade assays, analysis of mutant fusion loop epitopes, and antibody-mediated functional assays, including neutralization and FcγRIIa-mediated antibody-dependent enhancement assays mediated **(B)** JEV VLP **(C)** YFV NS1-reactive IgG levels over time **(D)** DENV1-4 VLP-reactive IgG levels at day 28. In panels **B, C, and D**, jitter plots show individual data points, with group classifications in the legends. Jitter plots display means with standard errors. P values for comparisons above the top bracket were obtained using the Kruskal-Wallis test with Dunn’s correction for multiple comparisons and are reported as means ± SEM. A p-value ≤0.05 was considered significant. The groups included JE-naïve (DV-CYD, orange, n=9), simultaneous administration (JE-IXIARO+DV-CYD, green, n=9), and JE IXIARO-priming (JE IXIARO→DV-CYD, blue, n=9). All MSD experiments were performed in a single replicate, as MSD offers high throughput, high sensitivity, and a wider dynamic range. Abbreviations: DV or DENV, dengue virus; JE or JEV, Japanese encephalitis virus; YFV, yellow fever virus; VLP, virus-like particle; FLE, fusion loop epitope; E, envelope protein; NS1, non-structural protein 1; Ig, immunoglobulin; D0, day 0; D28, day 28; D84, day 84; D196, day 196; D336, day 336; MSD, meso-scale discovery; ECL, electrochemiluminescence; CYD-TDV, chimeric yellow fever virus-derived tetravalent dengue virus vaccine.

### \Prior JEV vaccination with IXIARO establishes higher cross-reactive IgG reactivity to DENV VLPs

We first confirmed that JEV IXIARO and DENV CYD-TDV induced strong antibody responses by measuring IgG responses to both the JEV VLP and the YFV NS1 protein. We observed that JEV IXIARO-primed and simultaneously administered vaccinees had higher IgG levels against JEV VLP over time than JEV-naïve individuals (**Fig. 1B**), suggesting a robust JEV IXIARO immune response. Additionally, all individuals, regardless of JEV vaccine status, exhibited a robust, balanced YFV NS1 IgG response (**Fig. 1C**), confirming the immunogenicity of CYD-TDV.

We then examined whether the JEV IXIARO vaccination affects the overall DENV IgG response induced by CYD-TDV over time. By 28 days after the first CYD-TDV dose (D28), individuals who were primed with the inactivated JEV IXIARO vaccine showed significantly higher IgG levels binding to DENV1-4 VLPs than JEV-naïve individuals or those who received JEV IXIARO and DENV CYD-TDV simultaneously (**Fig. 1D**). The DENV1-4 IgG responses continued to grow throughout the CYD-TDV vaccination regimen in all groups, reaching a plateau after the third dose within each group (**Fig. S1A**). The breadth of this DENV IgG response varied across DENV serotypes and individuals at early time points (day 28) in JEV IXIARO-primed individuals, then expanded over time with subsequent CYD-TDV doses across all groups (**Fig. S1B**). We also examined the IgM response to DENV1-4 VLPs, finding similar levels across all groups that peaked by D28 (**Fig. S1C**) and then declined over time, regardless of additional CYD-TDV doses (**Fig. S1D**). Overall, these findings suggest that a robust, early cross-reactive DENV1-4 IgG response depends on prior flavivirus exposure rather than single or co-exposure.

### \Dengue vaccination with CYD-TDV preferentially recalls FLE-biased antibody responses following JEV IXIARO priming

To determine whether prior vaccination with inactivated JEV IXIARO affects dengue epitope-specific humoral immunity after CYD-TDV vaccination, we measured the proportion of antibodies that prevent 4G2 and 4E11 mAbs from binding FLE (4G2 FLE-like) (**Fig. 2A**) and the EDIII (4E11 EDIII-like) (**Fig 2A**) epitopes in the DENV envelope protein in post-vaccination sera, respectively, using a MSD-based blockade-of-binding (BOB) assay. Participants who received JEV IXIARO and DENV CYD-TDV simultaneously showed intermediate (day 28) 4G2 FLE-like responses at day 28, which were significantly lower than those in the sequential JEV IXIARO-primed group but similar to JEV-naïve individuals (**Fig. 2B**). All groups exhibited low or undetectable responses targeting the 4E11 EDIII epitope at day 28 (**Fig. 2C**). All groups showed an increase in 4G2 FLE-like antibody levels compared to 4E11 EDIII-like antibody after the two first doses of CYD-TDV (**Fig. S2A**). These findings indicate that inactivated JEV IXIARO vaccination is associated with a preexisting pool of antibodies targeting conserved DENV envelope fusion-loop epitopes, irrespective of subsequent CYD-TDV vaccination.

**Figure 2.**
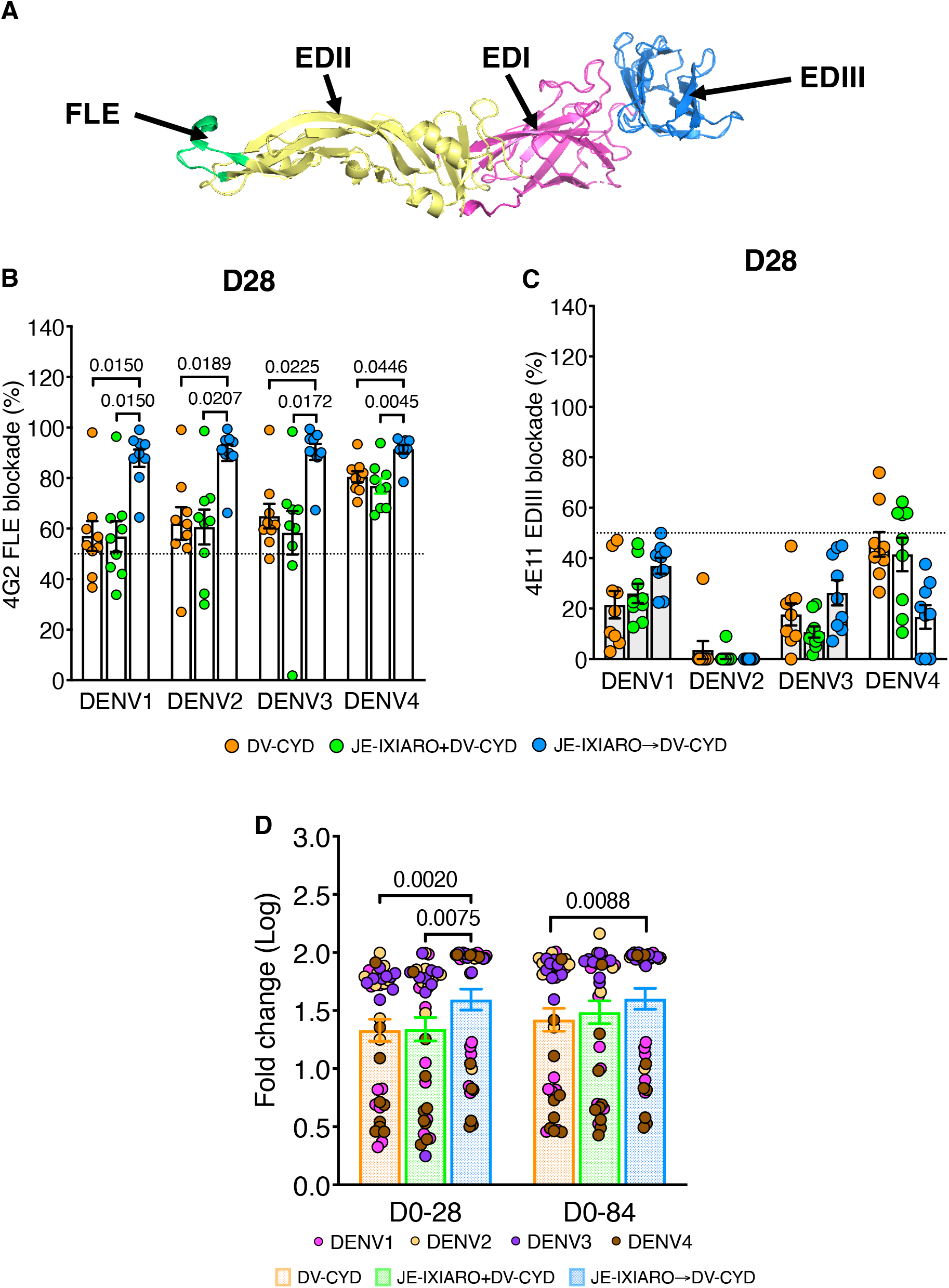
Prior JEV IXIARO vaccination induces fusion loop–reactive antibodies. **(A)** Structure of the DENV Envelope (E) protein monomer (PDB 1TG8)^55^, highlighting the fusion loop epitope (FLE, green) and the E domains. EDI is shown in magenta, EDII in yellow, and EDIII in marine blue. Levels of 4G2 FLE–like **(B)** and 4E11 EDIII-like **(C)** antibodies measured via epitope-blockade assay in JEV-naïve (DV-CYD) and IXIARO-vaccinated individuals before (JE-IXIARO→DV-CYD) or during (JE-IXIARO+DV-CYD) dengue CYD-TDV vaccination at day 28. **(D)** Changes in 4G2 FLE-like antibody levels from baseline to day 28. Bar plots show mean values with standard errors. P values above the top bracket are from Kruskal-Wallis tests with Dunn’s correction for multiple comparisons, reported as means ± SEM. A p-value of ≤0.05 indicates statistical significance. The groups included JE-naïve (DV-CYD, orange, n=9), simultaneous administration (JE-IXIARO+DV-CYD, green, n=9), and JE IXIARO-priming (JE IXIARO→DV-CYD, blue, n=9) groups. All MSD experiments were performed in a single replicate, as MSD offers high throughput, high sensitivity, and a wider dynamic range. Abbreviations: DV or DENV, dengue virus; JE or JEV, Japanese encephalitis virus; VLP, virus-like particle; Ig, immunoglobulin; D0, day 0; D28, day 28; D84, day 84; FLE, fusion loop epitope; E, envelope protein; EDIII, E domain III; MSD, meso-scale discovery; ECL, electrochemiluminescence; CYD-TDV, chimeric yellow fever virus-derived tetravalent dengue virus vaccine.

We then studied the long-term dynamics of 4G2 FLE-like antibody responses following immunization with the live-attenuated dengue CYD-TDV vaccine. The extent and timing of these responses varied significantly across vaccination platforms and schedules. Antibody levels were notably higher in individuals primed with JEV IXIARO, with the greatest difference observed on day 28 after the initial CYD-TDV dose (**Fig. S2B**). This expanded response persisted through day 84 after the second dose and declined after the third dose (day 196), eventually leveling off across all groups by day 336 (**Fig. S2B**). Analysis of fold-change relative to baseline confirmed that JEV IXIARO-primed individuals consistently showed a greater magnitude of FLE-reactive recall responses compared to simultaneous administration and JEV naïve individuals on days 28 and 84 (**Fig. 2D**). By day 336, this recall level decreased to around baseline level, as shown in **Figs. S2C and S2D**. These findings support the idea that JEV IXIARO-primed individuals preferentially recall preexisting 4G2 FLE-biased memory B cell responses after CYD-TDV vaccination. Additionally, the timing and platform of flavivirus vaccination play a role in determining the extent of fusion loop–biased immune recall.

### Fusion-loop–directed antibodies drive the dengue CYD-TDV vaccine response in JEV IXIARO-primed individuals

To directly assess the role of fusion loop–specific antibodies in the humoral response elicited by the CYD-TDV vaccine, we measured serum IgG binding to recombinant DENV-2 E protein dimers engineered to disrupt recognition by 4G2 FLE. We used both wild-type (WT) and 4G2 FLE-mutant DENV-2 E dimers (**Fig. 3A**) to distinguish antibodies that depend on FLE from those that do not. Individuals primed with JEV IXIARO showed a marked decrease in binding to 4G2 FLE-mutant DENV-2 E dimers compared to WT DENV-2 E (**Fig. S3A**). This difference persisted after the second CYD-TDV dose in JEV IXIARO-primed individuals (**Fig. S3B**). Conversely, sera from JEV-naïve individuals and from those administered simultaneously with IXIARO and CYD-TDV continued to bind significantly to FL-mut DENV-2 E dimers, suggesting that fusion loop disruption has less effect in these cases (**Figs. S3A, S3B**). These findings imply that co-administration reduces 4G2 FLE dominance by allowing the production of antibodies targeting non–fusion-loop epitopes.

**Figure 3.**
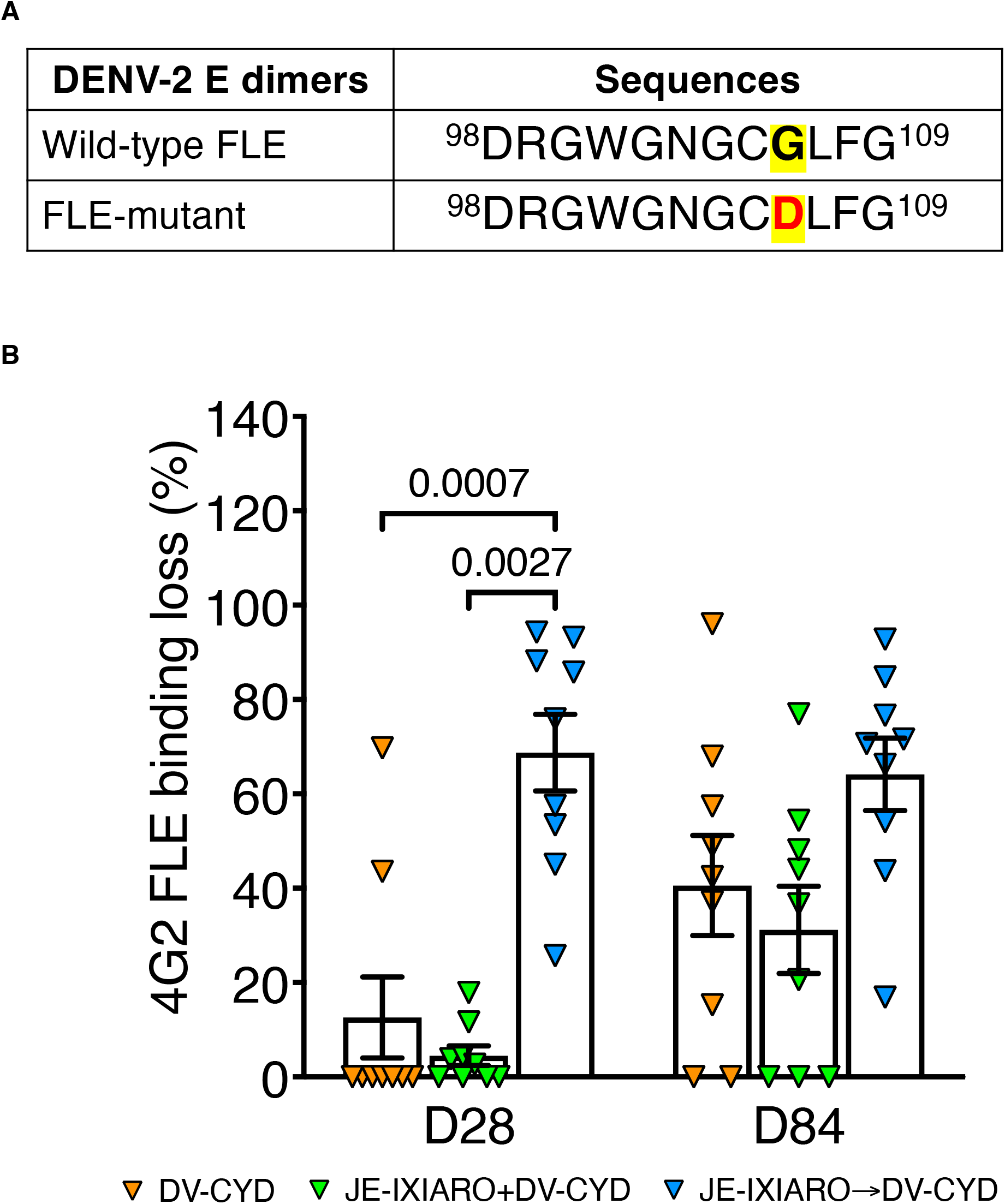
Memory immune recall of fusion loop–dominance influences early CYD-TDV vaccination response. **(A)** Sequences of wild-type and Fusion loop epitope-mutant DENV-2 E-dimers. Both FLE regions span amino acids 98-109 of the E protein^56^. The FLE-mutant has a mutation at position 106, highlighted in red and marked in yellow. **(B)** Sequential priming with IXIARO results in higher initial FLE-reactive antibody levels and faster recall of 4G2 FLE–focused responses after dengue CYD-TDV vaccination. In contrast, simultaneous priming diminishes 4G2 FLE dominance. Individual data points are plotted, with group labels in the legends. Fusion loop dominance was calculated as the percentage (%) loss in 4G2 FLE binding to FLE-mut compared to WT E dimers, using the formula: ((DENV-2 WT E dimer – DENV FLE-mut E dimer)/DENV-2 WT E dimer) × 100. Percentages below 0 were set to 0. P values above the top bracket are from Kruskal-Wallis tests with Dunn’s correction for multiple comparisons, expressed as means ± SEM. A p-value of ≤0.05 indicates statistical significance. Groups include JE-naïve (DV-CYD, orange, n=9), simultaneous administration (JE-IXIARO+DV-CYD, green, n=9), and JE IXIARO-priming (JE IXIARO→DV-CYD, blue, n=9). All MSD experiments were performed in a single replicate because MSD offers high throughput, high sensitivity, and a wider dynamic range. Abbreviations: FLE, fusion loop epitope; DV or DENV, dengue virus; JE or JEV, Japanese encephalitis virus; VLP; Ig, immunoglobulin; D28, day 28; D84, day 84; MSD, meso-scale discovery; CYD-TDV, chimeric yellow fever virus-derived tetravalent dengue virus vaccine.

Analysis of dominance on the 4G2 FLE, measured by the fractional reduction in binding to 4G2 FLE-mut compared to WT DENV-2 E dimers, showed significantly stronger 4G2 FLE dominance in JEV IXIARO-primed individuals at day 28, with this effect persisting at day 84 (**Fig. 3B**). This bias toward 4G2 FLE is also evident when assessing the magnitude of IgG-reactive 4G2 FLE antibodies (**Fig. S3C**). Together, these findings demonstrate that prior JEV IXIARO vaccination shapes the CYD-TDV antibody response toward fusion-loop–dependent epitopes and that this imprinting is dominant during early recall. The use of a fusion loop mutant E dimer provides direct evidence that fusion loop–specific antibodies constitute a significant component of the early CYD-TDV –elicited antibody response in JEV-primed individuals.

### FLE-biased antibody responses are associated with cross-Flavivirus broad neutralization

To measure the functional antibody response following a 4G2 FLE-biased recall during sequential Flavivirus exposure, we measured neutralizing antibody titers against DENV1-4 using the PRNT50 assay. After the initial CYD-TDV dose, all groups exhibited moderate neutralizing capacity against all four DENV serotypes (**Fig. 4A**), suggesting that individuals primed with JEV IXIARO had DENV neutralizing titers comparable to those of co-administered and JEV-naïve individuals after the first dose of CYD-TDV. While the overall mean DENV neutralizing titers were similar across groups, titers varied by serotype and group, with neutralization being consistently lowest against DENV1 in all groups over time (**Fig. S4A**).

**Figure 4.**
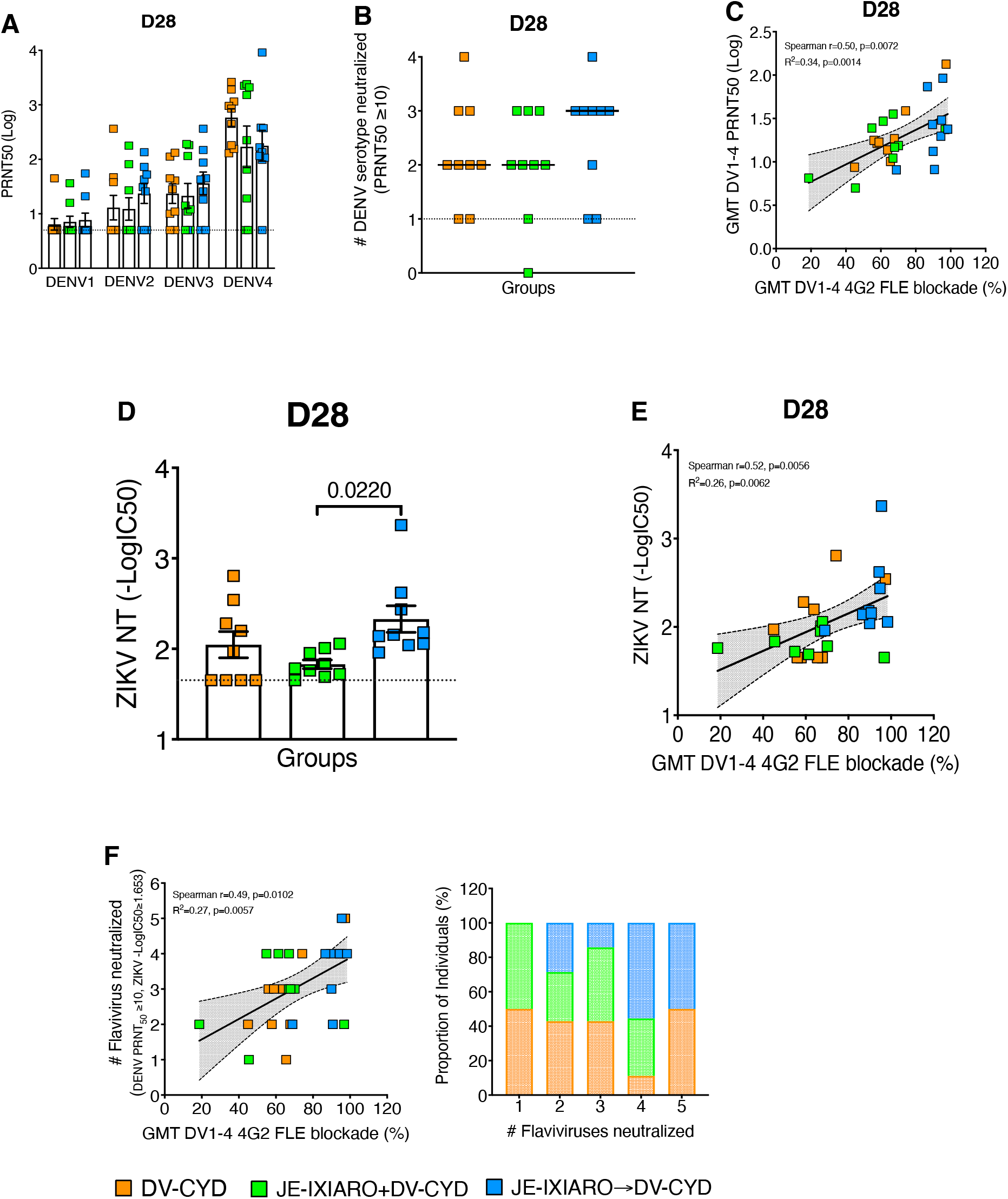
Fusion loop imprinting promotes broad but low-potency neutralization. **(A, D)** Neutralization titers against DENV serotypes 1–4 **(A)** and Zika virus (ZIKV) **(D)** were measured on day 28 after CYD-TDV vaccination in the following individual groups: JEV-naïve, simultaneous administration, and JE IXIARO priming. P values above the top bracket are from Kruskal-Wallis tests with Dunn’s correction for multiple comparisons and are shown as means ± SEM. A p-value of ≤0.05 indicates statistical significance. **(C, E)** Scatterplots show the correlation between the geometric mean (GMT) of DENV1-4 4G2 FLE-like antibody levels and either the GMT of DENV1-4 **(C)** or the level of ZIKV-neutralizing antibody **(E)**. **(B, F)** Neutralization breadth reflects the number of DENV serotypes 1-4 **(B)** or flaviviruses, including DENV1-4 and ZIKV **(F)**, neutralized above the threshold for each individual. Fusion loop–biased responses in JEV IXIARO-primed individuals are linked to increased neutralization breadth but only modest neutralization potency. Data are presented as geometric mean titers, with individual data points included. Groups include JE-naïve (DV-CYD, orange, n=9), simultaneous administration (JE-IXIARO+DV-CYD, green, n=9), and JE IXIARO-priming (JE IXIARO→DV-CYD, blue, n=9). All MSD experiments were conducted in a single replicate because of MSD’s high throughput, sensitivity, and wide dynamic range, whereas all neutralization experiments were performed in technical duplicates. Abbreviations: DV or DENV, dengue virus; JE, Japanese encephalitis; VLP, virus-like particle; ZIKV, Zika virus; NT, neutralization titer; PRNT, plaque reduction neutralization test; GMT, geometric mean titer; FLE, fusion loop epitope; Ig, immunoglobulin; MSD, meso-scale discovery; ECL, electrochemiluminescence; CYD-TDV, chimeric yellow fever virus-derived tetravalent dengue virus vaccine.

We then explored how 4G2 FLE-biased JEV IXIARO priming relates to the breadth of DENV neutralization, measured by the number of serotypes neutralized above a PRNT50 threshold of 10. Participants primed with JEV IXIARO showed a broader ability to neutralize 2-4 DENV serotypes after CYD-TDV vaccination, compared to the other groups (**Figs. 4B, S4B-S4C**). Additionally, the overall geometric mean neutralization titers (GMTs) for DENV1-4 were significantly correlated with the early-stage 4G2 FLE-like antibody response (day 28) in the CYD-TDV response (**Fig. 4C**). When stratified by vaccine history, this correlation was not observed in JEV IXIARO-immunized individuals compared with JEV-naïve subjects (**Fig. S4D**). The levels of 4G2 FLE-like antibodies by serotype display variable patterns, from no correlation to moderate correlation with neutralization potencies across all four DENV serotypes, regardless of prior flavivirus vaccination (**Fig. S4E**). These results suggest that while 4G2 FLE-biased antibody recall broadens DENV binding, it does not linearly enhance neutralizing potency in JEV IXIARO-primed participants.

We further examined whether cross-flavivirus neutralization driven by 4G2 FLE-biased JEV immune imprinting extends beyond the DENV serocomplex. Using a ZIKV reporter virus particle (RVP) flow-based neutralization (NT) assay, we measured neutralization activity against ZIKV. At day 28, individuals primed with JEV IXIARO exhibited higher ZIKV neutralizing titers than JEV-naïve individuals. However, this difference was not statistically significant. Conversely, those vaccinated simultaneously with JEV IXIARO and DENV CYD-TDV showed lower ZIKV neutralization titers (**Fig. 4C**). The overall level of 4G2 FLE-like antibodies correlates with ZIKV neutralization titers (**Fig. 4D**). This suggests that co-vaccination promotes the recruitment of naive B cells targeting diverse epitopes, reducing the dominance of 4G2 FLE-like antibodies and resulting in a more balanced antibody response.

When quantifying cross-flavivirus neutralizing antibodies by the number of flaviviruses neutralized above a set threshold, the overall 4G2 FLE-like antibody was significantly linked to a higher number of neutralized flaviviruses. Only a small fraction of individuals in the JEV-naïve and simultaneous groups neutralized four to five flaviviruses. In contrast, the JEV IXIARO-primed group had a higher proportion of individuals who neutralized four to five flaviviruses (**Figs. 4E, S4F**). These findings suggest that 4G2 FLE-biased responses triggered by JEV IXIARO priming did not affect the strength of flavivirus neutralization but did contribute to broad, low-potency cross-flavivirus neutralization, which aligns with the conserved nature of FLE across flaviviruses.

### Fusion loop-biased imprinting predicts FcγRIIa-mediated *in vitro* antibody-dependent enhancement

Since antibody function extends beyond neutralization, we also investigated Fc receptor–mediated antibody-dependent enhancement (ADE) using FcγRIIa-expressing K562 cells *in vitro*. Individuals primed with JEV IXIARO showed higher ADE signals at intermediate serum dilutions, particularly at the early post-vaccination point (day 28) (**Fig. S5A**). Analyzing the peak enhancement titer (PET) and the overall enhancement (area under the curve, ADE AUC) demonstrated significantly increased ADE activity in the group primed with JEV IXIARO compared to both JEV-naïve and simultaneously administered groups (**Figs. 5A, 5B**). Consistent with decreased fusion loop dominance, sera from individuals vaccinated simultaneously with JEV IXIARO exhibited lower enhancement signals than those from the sequentially vaccinated group (**Figs. 5A, 5B**). Although DENV1-4 serotype PRNT50 titers showed weak, inconsistent correlations with ADE magnitude across all groups (**Fig. S5B**), overall neutralization titers did not correlate with enhancement levels (**Fig. 5C**), suggesting that *in vitro* ADE is not solely governed by neutralization strength. Next, we evaluated how the 4G2 FLE-like antibody and vaccination history, specifically, the platform and timing, predict FcγRIIa-mediated *in vitro* ADE. Correlation analysis revealed a moderate association between 4G2 FLE-like Ab levels and FcγRIIa-mediated enhancement at early time points (day 28) in individuals primed with JEV IXIARO. In contrast, this association was weaker in JEV-naïve participants and was not observed in those vaccinated simultaneously (**Fig. 5D**).

**Figure 5.**
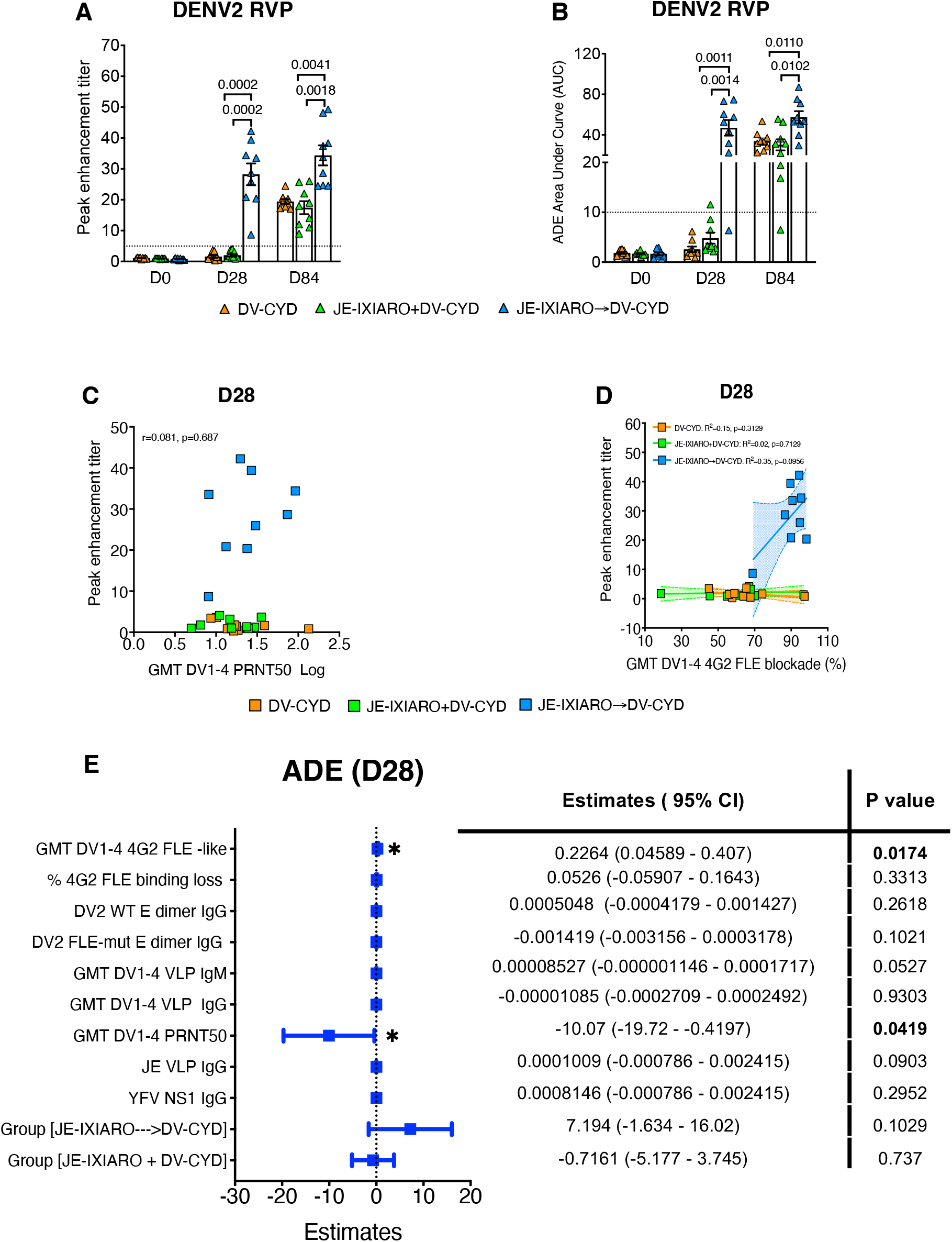
Fusion loop–biased imprinting is associated with FcγRIIa-mediated antibody enhancement at day 28. **(A. B)** Quantification of ADE activity is shown through peak enhancement titer (PET) and the area under the enhancement curve (AUC) calculated from ADE curves. **(C)** There is a correlation between the GMTs of DENV1-4 neutralization and ADE activity (PET) after dengue CYD-TDV vaccination. **(D)** A linear relationship was observed between the GMTs of DENV1-4 4G2 FLE-like levels and ADE activity at day 28, as shown by correlation and simple regression analysis. (E) An adjusted multivariable regression identified independent predictors of FcγRIIa-mediated ADE at day 28; notably, the 4G2 FLE bias remained independently associated with ADE after adjusting for covariates, including vaccine history, neutralization titers, and total DENV-binding IgG. Standardized coefficients with 95% confidence intervals are provided. Groups include JE-naïve (DV-CYD, orange, n=9), simultaneous administration (JE-IXIARO+DV-CYD, green, n=9), and JE IXIARO-priming (JE IXIARO→DV-CYD, blue, n=9). All MSD experiments were conducted in a single replicate due to MSD’s high throughput, sensitivity, and wider dynamic range, while all ADE experiments were performed in technical duplicates. Abbreviations: DV or DENV, dengue virus; JE, Japanese encephalitis virus; YFV, yellow fever virus; GMT, geometric mean titer; PRNT, plaque reduction neutralization test; FLE, fusion loop epitope; mut, mutant; NS1, non-structural protein 1; D28, day 28; VLP, virus-like particle; RVP, reporter virus particle; Ig, immunoglobulin; ADE, antibody-dependent enhancement; MSD, meso-scale discovery; CYD-TDV, chimeric yellow fever virus-derived tetravalent dengue virus vaccine.

Furthermore, we performed univariate and multivariable analyses on day 28, the period of peak recall, to determine whether 4G2 FLE–biased antibody responses influence FcγRIIa-mediated ADE following dengue CYD-TDV vaccination. In univariable analyses, the level of 4G2 FLE-like antibodies and their dominance in fusion loop dependence, assessed by the percentage loss of 4G2 FLE-mutant binding, were both positively associated with the magnitude of ADE (**Fig S5C**).

In multivariable models that included the levels of 4G2 FLE-like Ab, 4G2 FLE dominance, DENV-2 wild-type, and fusion loop-mutant E dimer-reactive IgGs, the geometric means of DENV1-4 neutralizing titers, total DENV1-4 binding IgM, and total DENV1-4-reactive IgGs identified 4G2 FLE-like Ab and 4G2 FLE dominance as significant predictors of ADE activity (PET) at day 28. The geometric mean of DENV1-4 PRNT50 and other predictors did not contribute to the model (**Fig. S5D**). These findings suggest that epitope specificity, rather than neutralization potency, primarily influences early FcγRIIa-mediated ADE *in vitro* and support a model in which fusion loop–biased imprinting drives ADE and antibody-mediated host–pathogen interactions through Fc receptor engagement. Adjusting for vaccination history (JEV-naïve, simultaneous administration, JEV IXIARO priming) and for the levels of JEV VLP- and YFV NS1-reactive IgGs reduced but did not eliminate the link between 4G2 FLE-like Ab and ADE (**Fig 5E**). This link reinforces the notion that the 4G2 FLE bias functions as a mechanistic factor behind group-level differences in FcγRIIa-mediated ADE.

### Full, three-dose live-attenuated CYD-TDV vaccination series overcomes 4G2 FLE–biased immune imprinting

To understand how antigen dose and replication capacity influence 4G2 FLE–biased immune imprinting, we studied antibody responses in a subgroup of individuals who were initially primed with the JEV IXIARO vaccine and subsequently received all three doses of the live-attenuated CYD-TDV vaccine series. Compared with the response after a single CYD-TDV dose (day 28), the subsequent dose gradually reduced the dominance of 4G2 FLE–like responses and altered the antibody profile in JEV-IXIARO-primed individuals. By days 196 and 336, these profiles resembled those of JEV-naïve and simultaneously administered individuals, respectively (**Figs. 6A, S6A**). Consequently, the 4E11 EDIII titers moderately rose following a complete CYD-TDV vaccination series, matching the levels of 4G2 FLE-like antibodies at day 336 in all JE IXIARO-immunized individuals (**Figs. 6B, S6B, and S6C**). Structural epitope analysis using the FLE-mut E-dimer assay revealed increased antibody binding to FLE-mut DENV-2 E dimers, resulting in the elimination of the gap between WT and FLE-mut E-dimer binding in JEV IXIARO-primed participants after receiving the full CYD-TDV series (**Fig. 6C**). Together, these data indicate that administering the full, three-dose series of CYD-TDV overcomes the JEV IXIARO priming effect by eliciting a new B-cell response that produces antibodies targeting non–fusion-loop epitopes, such as EDIII and the E-dimer epitope (EDE). Along with this change in epitope recognition, there was also a reduction in FcγRIIa-mediated antibody-dependent activity, even after priming with JEV IXIARO (**Figs. 6D, S6D, and S6E**). Overall, these findings demonstrate that the 4G2 FLE–biased immune imprinting is not fixed; it can be influenced by higher and repeated antigen exposures. This observation underscores the importance of antigen dose and the vaccine platform as additional determinants of epitope dominance and antibody responses.

**Figure 6.**
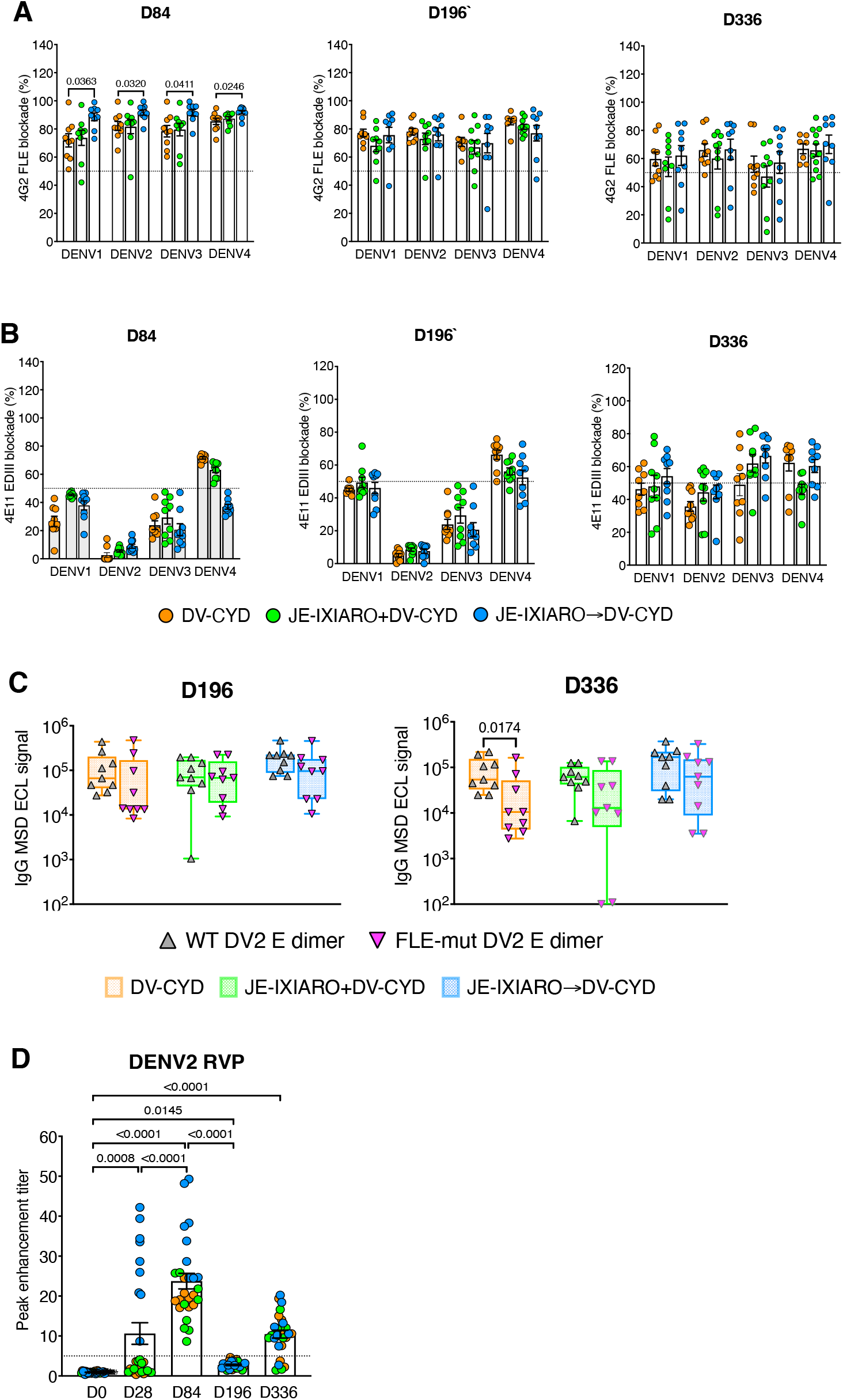
Full-three dose series of live-attenuated dengue CYD-TDV vaccination reduces fusion loop imprinting. Comparison of uptake between partial (day 84) and complete (days 196 and 336) dose series of live-attenuated dengue CYD vaccination across DENV serotypes. **(A, B)** Responses to 4G2 FLE– like and 4E11 EDIII-like antibodies. **(C)** Binding activity to wild-type (WT) and fusion loop mutant (FLE-mut) envelope dimers at days 196 and 336, showing increased binding retention to FLE-mut dimers after the full dose. **(D)** Long-term FcγRIIa-mediated ADE activity over time and within the dengue CYD-TDV vaccination schedule. A full dose of dengue CYD-TDV enhances antigenic stimulation, reduces 4G2 FLE–biased imprinting, and decreases FcγRIIa-mediated antibody enhancement. Groups include JE-naïve (DV-CYD, orange, n=9), simultaneous administration (JE-IXIARO+DV-CYD, green, n=9), and JE IXIARO-priming (JE IXIARO→DV-CYD, blue, n=9). All MSD experiments were conducted in a single replicate because of MSD’s high throughput, sensitivity, and wide dynamic range, whereas all ADE experiments were performed in technical duplicates. Abbreviations: DV or DENV, dengue virus; JE or JEV, Japanese encephalitis virus; YFV, yellow fever virus; VLP, virus-like particle; RVP, reporter virus particle; E, envelope protein; EDIII, E domain III; FLE, fusion loop epitope; mut, mutant; WT, wild-type; D28, day 28; D84, day 84; D196, day 196; Ig, immunoglobulin; MSD, meso-scale discovery; ECL, electrochemiluminescence; CYD-TDV, chimeric yellow fever virus-derived tetravalent dengue virus vaccine.

## DISCUSSION

Vaccination remains the most effective method to reduce the impact of flavivirus diseases, including JEV and DENV. JEV IXIARO is primarily used by travelers and residents in JE-endemic areas, including parts of Asia where other flaviviruses, such as DENV and ZIKV, are also prevalent. In these regions, live-attenuated DENV vaccines such as CYD-TDV and Qdenga have been approved and are in use^40^ affecting vaccine immunogenicity, efficacy, disease outcomes, and overall infection force. This raises important questions about how preexisting immunity influences the response to subsequent vaccination or infection with heterologous flaviviruses, a phenomenon known as immune imprinting. This occurs when the immune system, after an initial infection with viruses like DENV, ZIKV, or JEV, responds differently to subsequent related infections, often favoring a memory response to the first virus ^41^. In flavivirus contexts, immune imprinting has been extensively studied in both natural infection and vaccination^7,8,42–47^.

While imprinting is typically associated with changes in antibody binding or neutralization, our study further investigated how it can influence epitope-level binding, neutralization, and Fc receptor– mediated enhancement during sequential flavivirus vaccination. This expands our understanding of how immunity from multiple vaccines develops, especially in regions where flaviviruses circulate simultaneously, and routine vaccination against one or more is common. Importantly, we examined how prior vaccination with a purified inactivated vaccine (JEV IXIARO) affects antibody responses and their functions after live-attenuated vaccination (CYD-TDV).

Our study indicated that by day 28, participants who received the JEV IXIARO vaccine developed significantly higher levels of cross-reactive IgG antibodies against DENV than the JEV-naïve group or those who received the vaccines simultaneously. This aligns with findings from other flaviviruses, such as TBEV and YFV, where, by day 28 post-vaccination, individuals preimmunized with TBEV exhibit higher cross-reactive IgG antibody levels to YFV than TBEV-naïve individuals^7^. These data suggest that prior immunity from inactivated JEV IXIARO vaccination redirects the dengue CYD-TDV vaccine response toward conserved E protein epitopes. It remains to be seen whether this effect is specific to the inactivated JEV vaccine or also applies to JEV live-attenuated vaccines. Using a FLE-mut E dimer binding assay to evaluate antibody targeting of the conserved 4G2 FLE epitope, we found that the 4G2 FLE binding antibody is predominant and makes up a large part of the CYD-TDV vaccine response following sequential JEV IXIARO and CYD-TDV vaccination. This analysis provides insights beyond peptide-based assays, indicating that 4G2 FLE-like imprinting occurs at the epitope level rather than through cross-reactivity. It also highlights the preferential recall of cross-reactive memory B cell responses, which could affect antibody function.

This study enhances traditional immune imprinting models by connecting preferred epitopes to functional antibody responses. FLE–directed antibodies are known for their broad cross-reactivity but are only weakly neutralizing^48^. Our results show that individuals primed with JEV IXIARO exhibit broad yet low-potency neutralization across dengue serotypes and Zika virus, indicating that 4G2 FLE-biased responses—induced by heterologous sequential flavivirus vaccination—are linked to neutralization breadth. This aligns with recent studies, such as Hattakam *et al*., who showed that sequential DENV exposure produces potent cross-neutralizing antibodies against DENV and ZIKV^5^, and Salem *et al*., who found that antibodies from DENV patients with prior JEV exposure are broadly neutralizing against ZIKV

^8^. Additionally, a novel cross-reactive monoclonal antibody, mAb 2A10G6, targeting a newly identified epitope within the highly conserved flavivirus FLE, has demonstrated broad neutralizing activity against several flaviviruses, including DENV1-4, JEV, TBEV, and WNV^49^. To address the gap between inactivated vaccination priming and neutralization potency, we measured DENV-reactive IgM antibodies. Participants who were JEV-naïve or vaccinated simultaneously showed IgM levels comparable to those of JEV IXIARO-primed individuals, suggesting that DENV IgM may contribute more to DENV neutralization than IgG at the early stage of antibody response. This is consistent with Santos-Peral *et al*.’s report that preimmunization with inactivated TBEV did not affect live-attenuated YFV vaccine neutralization, which was primarily IgM-dependent.

Our research shows that the 4G2 FLE–biased antibody, induced by JEV IXIARO vaccination, predicts FcγRIIa-mediated antibody-dependent enhancement (ADE). This result aligns with recent research by Santos-Peral *et al*., who found that FLE-specific cross-reactive IgG from TBEV vaccination can promote DENV ADE^7^. This underscores epitope specificity as a key determinant of antibody effectiveness. It also highlights the distinction between neutralization and Fc-mediated enhancement, underscoring the importance of assessing antibody responses across multiple functional properties. Although ADE provides a sensitive indicator of antibody quality beyond what neutralization tests reveal, especially for flaviviruses, where antibody level, affinity, and epitope specificity collectively impact functional outcomes. We view FcγRIIa-mediated enhancement as an *in vitro* functional characteristic rather than a direct predictor of disease in humans, as early research indicated that the JEV vaccine elicits cross-reactive antibodies targeting conserved epitopes, thereby affecting both clinical and functional outcomes of DENV^38^. Anderson *et al*. initially reported more symptomatic dengue cases among JE-vaccinated individuals compared to unvaccinated ones ^6^. Later, Saito *et al*. found that approximately 45% of JEV-vaccinated individuals develop ADE-capable antibodies upon DENV infection. The JE vaccine was also linked to a higher risk of severe dengue during initial DENV infection ^43^. More studies are essential to better understand whether the FLE antibody imprinting induced by JEV vaccination confers protective cross-reactive immunity or increases disease severity via antibody-dependent enhancement in subsequent DENV vaccination or infection.

The 4G2 FLE imprinting was not permanent. Higher antigenic stimulation from the full three-dose live-attenuated dengue vaccine series decreased the dominance of 4G2 FLE, increased the recruitment of antibodies targeting non–fusion loop epitopes, and shifted antibody function toward less FcγRIIa-mediated enhancement. These results support a model in which epitope dominance depends on the interplay between memory recall and new B cell recruitment, and is influenced by the vaccine platform, timing, and antigen dose.

This study has certain limitations. First, Fc receptor–dependent antibody activity was assessed using *in vitro* assays with FcγRIIa-expressing cells, and the *in vivo* impact of 4G2 FLE imprinting remains unexplored. Second, although the FLE mutant E-dimer assay indicated 4G2 FLE dominance, it does not provide details about the clonal composition of the antibody response. Future research combining single-cell B cell profiling with monoclonal antibody isolation will be crucial for understanding the cellular and clonal basis of this imprinting. Third, longitudinal studies linking epitope-specific antibody function to clinical outcomes will be important to evaluate the translational relevance of FLE imprinting in vaccinated populations. Lastly, since both this and previous studies involved Flavivirus-naïve participants, it’s unclear whether this phenomenon is limited to vaccine-induced immunity rather than natural infection in such individuals. Additional research in flavivirus-endemic areas, where viruses circulate together, people experience repeated exposures, and vaccination programs often involve sequential heterologous vaccines, is necessary to understand how immune history influences antibody quality. Extending these analyses to other flavivirus vaccines and platforms will determine if these findings are widely applicable. Despite these limitations, a notable strength is that prior vaccination with the inactivated vaccine (JEV IXIARO) shapes the antibody response to live-attenuated dengue CYD-TDV vaccination toward conserved fusion-loop epitopes, thereby promoting FLE-immune imprinting. This immune imprinting targeting conserved FLE epitopes may be a general principle through which flavivirus immune history shapes humoral responses to related antigens. This mechanism likely extends beyond flavivirus, influencing antibody responses after vaccination or infection with other antigenically related viruses. Additionally, our results provide a framework for understanding how different vaccination strategies affect antibody quality and activity, which is important for developing and evaluating flavivirus vaccines in populations with diverse immune backgrounds.

## METHODS

### Trial design, subjects, and sampling

This study extends our previously published phase II randomized, open-label trial conducted at the State University of New York (SUNY) Upstate Medical University^39^. It was a longitudinal observational study examining antibody responses following dengue vaccination in 45 healthy adults aged 18 to 45 years with no prior flavivirus exposure. Participants received either the inactivated JEV vaccine (IXIARO) or the live-attenuated dengue vaccine (CYD-TDV), in accordance with national or institutional guidelines. They were categorized into three groups based on vaccination sequence: JEV-naïve (n=15), sequential priming (IXIARO followed by CYD-TDV, n=15), or simultaneous priming (IXIARO and CYD-TDV administered within the same immunization window, n=15). Blood samples were collected at baseline (day 0, prior to dengue CYD-TDV vaccination) and longitudinally at days 28, 84, 196, and 336 following dengue CYD-TDV vaccination. Serum was isolated by centrifugation and stored at −80⍰°C until analysis. All participants provided written informed consent. The trial was approved by the institutional review boards at SUNY Upstate and registered on ClinicalTrials.gov (NCT01943825). A subset of individuals (n=27), including JEV-naïve (n=9), simultaneously administered (n=9), and JE IXIARO-primed (n=9), who completed all visits, was selected and analyzed in the current study.

### Vaccines

The inactivated JEV vaccine used in this study, IXIARO (Valneva), was administered at 1-month intervals. Dengue vaccination involved the live-attenuated tetravalent vaccine CYD-TDV (Sanofi Pasteur). The full-dose CYD-TDV was administered at 0, 2, and 6 months, according to the accelerated schedule outlined in the study protocols. All vaccine lot numbers and administration schedules were documented for each participant.

### Cells, recombinant E proteins, and VLPs

K562 cells were cultured at 37°C with 5% CO_2_ in IMDM medium (Gibco, #12440046), supplemented with 10% FBS, 1% L-glutamine, and 10% penicillin/streptomycin. Conversely, the RAJI-DCSIGN+ cell line, kindly provided by the NIAID Biodefense and Emerging Infectious Diseases (BEID) program, was maintained in R10 medium, consisting of RPMI (Gibco, #10-040-CM) supplemented with 10% FBS, 1% L-glutamine, and 1% penicillin/streptomycin. Recombinant stabilized DENV-2 (construct SC-14) E dimer proteins, derived from World Health Organization reference strain sequences for DENV-2 (strain 16681, amino acids 1–394), were produced by the University of North Carolina Protein Core as previously described^30,31^. A recombinant stabilized DENV-2 (strain 16681) E dimer protein (construct SC.10) containing a fusion loop epitope mutation at position 106 (G106D) was also developed and produced at the University of North Carolina, Chapel Hill, as previously described^31^. DENV serotypes 1–4, JEV virus-like particles (VLPs), and YFV non-structural-1 (NS1) recombinant antigens were purchased from the Native Antigen Company (Oxford, UK).

### Multiplex MSD-based binding assays for DENV1-4, JEV VLPs, and YFV NS1

To measure plasma IgG and IgM binding to DENV1-4, JEV, and YFV VLPs, we used the multiplex Meso Scale Discovery (MSD) Electrochemiluminescence (ECL) U-plex Development Pack (cat. # K15232N) as previously described^50^. DENV1-4, JEV VLPs, and YFV NS1 antigens were biotinylated separately using an EZ-Link SULFO-NHS-LC-Biotinylation Kit (ThermoScientific, cat# 21335) and then mixed at a 1:1 ratio. MSD plates coated with biotinylated VLPs of DENV1-4, JEV, and YFV NS1, conjugated to MSD linker proteins, were incubated for 1 hour at room temperature (RT) with shaking at 700 rpm. Plates were washed with wash buffer containing 0.05% Tween 20 (P7949, Sigma-Aldrich) and 1xPBS (PBS-T), then incubated with blocking buffer (1xPBS supplemented with 5% Bovine serum Albumin [BSA, 232100, BD Difco]) for 1 hour at RT with shaking at 700 rpm. After washing the plate with PBS-T, human plasmas diluted 1:5000 in MSD diluent (MSD, cat. #R50AA-3) were added, and the plate was incubated for 2 hours at RT with shaking at 700 rpm. Plates were then washed as described above and incubated with MSD GOLD SULFO-TAG Anti-human IgG (MSD, cat # D21ADF-3) or Anti-human IgM (MSD, cat # D21ADD-3) secondary antibody for 1 hour at RT with shaking at 700 rpm. Finally, the MSD plates were washed with the PBS-T, and the MSD Gold Read Buffer B (MSD, cat# R60AM-2) was added to the plate. The ECL signals were read as MSD readout values on an MSD instrument. DENV-naïve human plasma served as a negative control, and DENV polyclonal human plasma (SeraCare) served as a positive control. All plasmas with an MSD value greater than 100 were considered to have binding activity. All experiments were performed in a single replicate, as MSD offers high throughput, high sensitivity, and a wider dynamic range.

### Multiplex MSD-based assays for DENV-2 E dimer binding

DENV-2 wild-type (WT) E dimers and fusion loop epitope mutant (FLE-mut) E dimers with defined structures were used to assess FLE-specific antibody binding. The FLE-mut E dimers carry a specific amino acid change (G106) that selectively reduces the accessibility of the fusion loop epitope while preserving the overall E-dimer structure. MSD plates were coated with biotinylated WT or FLE-mut E dimers attached to MSD connectors and blocked as instructed. Plasma samples, diluted 1:5000, were added and incubated for 1 hour at room temperature with shaking. Bound antibodies were detected using MSD-conjugated anti-human IgG, and binding levels were measured with an MSD ECL value over 100. Fusion loop dominance was calculated as the percentage (%) loss in 4G2 FLE binding to FLE-mut compared to WT E dimers, using the formula: ((DENV-2 WT E dimer – DENV FLE-mut E dimer)/DENV-2 WT E dimer) × 100. Percentages below 0 were set to 0. All experiments were performed in a single replicate, as MSD offers high throughput, high sensitivity, and a wider dynamic range.

### Multiplex MSD-based epitope Blockade of binding (BOB)

This assay, derived from the previously described BOB ELISA^33,51,52^, was optimized on the multiplex MSD ECL platform in accordance with the manufacturer’s guidelines. FLE-specific IgG responses were measured by testing how human plasma blocks the binding of monoclonal antibodies 4G2 or 4E11 to the conserved fusion loop or E domain III (EDIII) epitope of recombinant DENV1-4 E protein, using an MSD ECL U-plex Development Pack. DENV1-4 virus-like particles (VLPs) were separately biotinylated with an EZ-Link SULFO-NHS-LC-Biotinylation Kit (ThermoScientific, cat# 21435) and mixed evenly. These biotinylated VLPs were coated onto MSD 96-well plates via conjugation to MSD linker proteins, then incubated for 1 hour at room temperature with shaking at 700 rpm. After washing with PBS-T and blocking with the specified buffer for 1 hour at room temperature with shaking, the plates were washed again. Human plasma samples and controls (from DENV-immune SeraCare and naïve individuals) shown in **Fig. S7A-B**, diluted 1:10 in MSD diluent 100 (cat# R50AA-3), were added to block FLE or EDIII epitopes and incubated for 2 hours at room temperature with shaking. The plates were washed and incubated with in-house 4G2 (ENVIGO, PUR0001) or 4E11 (Genscript) mAbs labeled with MSD GOLD SULFO-TAG NHS-Ester reagent (cat. # 31AA-1) for 1 hour at room temperature with shaking. Following a final wash, MSD Gold Read Buffer B (cat# R60AM-2) was added for detection. The ECL signal was read with an MSD instrument, and the percentage inhibition of 4G2 FLE or 4E11 EDIII binding was calculated relative to the no-plasma control using: % inhibition= 100 – ((ECL sample / ECL mock control) * 100). The titer of 4G2 FLE-like or 4E11 EDIII-like antibodies was defined as the proportion with ≥50% inhibition of binding. The absolute level of FLE-reactive IgG was determined by multiplying total IgG binding by the % inhibition. All experiments were conducted as single replicates, leveraging MSD’s high throughput, sensitivity, and broad dynamic range.

### Plaque Reduction Neutralization Test (PRNT) Assay

Neutralizing antibody titers against DENV1-4 were measured using a plaque reduction neutralization test (PRNT) as previously described and performed by Sanofi-Pasteur^39^. Serum samples were heat-inactivated and serially diluted before incubation with DENV1-4. Virus–serum mixtures were added to Vero cell monolayers and incubated under standard conditions. Plaques were visualized by immunostaining, and PRNT_50_ titers were calculated as the reciprocal serum dilution that reduced plaque counts by 50% relative to virus-only controls. Geometric mean titers were calculated for group comparisons.

### ZIKV RVP Generation and Neutralization Assay

ZIKV reporter virus particles (RVP) were produced by co-transfecting 293T cells with a plasmid encoding the ZIKV prME sequence (GenBank ID WEY06129.1), preceded by the WNV C gene sequence, alongside a WNVII-replicon plasmid, following the manufacturer’s Fugene-6 protocol. The transfected cells were incubated at 32°C for 48 hours to facilitate virion assembly, after which the media was replaced. Supernatants collected after 48 hours were centrifuged at 5,000g to remove cellular debris, filtered through 0.2 μm syringe filters, aliquoted, and stored at −80°C. For neutralization assays, plasma samples were serially diluted 4-fold, mixed equally with RVP, and incubated in duplicate wells of 96-well plates at 37°C for 1 hour. An equal volume of media containing 5 × 10^4^ RAJI-DCSIGN+ cells was then added to each well and incubated for 24 hours. Subsequently, the cells were washed twice with PBS + 2% FBS, fixed, and analyzed by flow cytometry to determine ZIKV-specific neutralization titers. All experiments were conducted in technical duplicates.

### DENV-2 RVPs-based K562 Antibody-dependent enhancement (ADE) assays

DENV-2 RVPs were produced as previously described ^53,54^. Briefly, RVPs were generated by co-transfecting 293T cells with a plasmid encoding the DENV-2 prME sequence (GenBank ID KM204118.1), preceded by the WNV C gene sequence, alongside a WNVII-replicon plasmid, following the manufacturer’s Fugene-6 protocol. The transfected cells were incubated at 32°C for 48 hours to facilitate virion assembly, after which the media was replaced. Supernatants collected after 48 hours were centrifuged at 5,000g to remove cellular debris, filtered through 0.2 μm syringe filters, aliquoted, and stored at −80°C. For ADE, FcγRIIa-mediated antibody function was assessed using ADE assays in K562 cells expressing FcγRIIa. Serial dilutions of serum were incubated with GFP-labelled DENV-2 RVPs before addition to K562 cells. After incubation, infection levels were quantified by GFP-expressing cells and flow cytometry. ADE activity was expressed as fold enhancement relative to virus-only controls. Area under the enhancement curve (AUC) and Peak enhancement titer (PET, the highest fold enhancement infection at a given sub-neutralizing dilution point) were calculated for each sample. All experiments were performed in technical duplicates.

### Multivariable analysis of ADE at day 28

To identify independent predictors of FcγRIIa-mediated ADE at day 28, multivariable linear regression models were developed. The main outcome was the peak enhancement titer. Predictor variables included 4G2 FLE-like, 4G2 FLE dominance, log_10_-transformed DENV PRNT_50_ titers, DENV1-4-binding IgM, total DENV1-4, JEV, and YFV-binding IgG. The vaccination group was incorporated as a covariate when relevant. Regression coefficients were standardized for effect-size comparison, and robust standard errors were used.

### Statistical analysis

Statistical analyses were conducted with GraphPad Prism. For group comparisons, nonparametric Kruskal-Wallis and Friedman tests with Dunn’s multiple comparisons were employed. Spearman’s rank correlation coefficient was used to assess correlations. Longitudinal data were analyzed using mixed-effects models. A P value of less than 0.05 was deemed statistically significant. No corrections were applied for multiple comparisons.

## Data Availability

All data produced in the present work are contained in the manuscript

## Data availability

All data supporting this study’s findings are included within the article and its supplementary files. No new reagents were created for this research. For additional information or to request resources and reagents, please contact the lead authors, Patrick I. Mpingabo at and Adam T. Waickman at.

## Code availability

No code available.

## Acknowledgement

We thank Dr. Mark Polhemus, Dr. Timothy Endy, and the Upstate Global Health Institute for designing and conducting the clinical trial that provided the samples. Our gratitude extends to the study participants and their families, as well as colleagues from the Department of Microbiology and Immunology at SUNY Upstate Medical University. The NIH HIV Reagent Program, Division of AIDS, NIAID, NIH provided the Raji DC-SIGN+ Cells (ARP-9945), contributed by Dr. Li Wu and Dr. Vineet N. KewalRaman. The 4G2 mAb was supplied under a Material Transfer Agreement from the Walter Reed Army Institute of Research (WRAIR). We sincerely thank Aravinda M. de Silva for sharing the DENV-2 E-dimers. Additionally, we appreciate Ted Pierson and Stephen Whitehead for their help with reporter-virus constructs and protocols. Our thanks also go to all members of the Waickman group at SUNY Upstate Medical University for their valuable input and contributions. The material has been reviewed and approved by the Walter Reed Army Institute of Research, with no objections to its presentation or publication. The views expressed are those of the author and do not reflect official positions of the Department of the Army or Department of Defense. The investigators have complied with AR 70–25 regulations regarding the protection of human subjects.

## Contribution

A.T.W., S.J.T., and K.B.A. acquired funding and supervised the current project. P.I.M., A.T.W., and S.J.T. conceived the current study. P.I.M., A.T.W., and G.D.G. contributed to the development of the study model. P.I.M. and A.T.W. designed all the experiments. J.Q.L. helped design MSD-based assays. P.I.M. performed the MSD-based epitope blockade of binding assay, DENV1-4, JEV, YFV, and E dimer binding assays, and the DENV-2 RVP-based K562 ADE assay. E.I.A. generated ZIKV RVPs and performed neutralization assay. M.H. produced DENV-2 RVPs. H.F. managed data curation and visualization tasks for the clinical trial. L.A.W. provided local oversight of the clinical trial. P.I.M. conducted formal analysis, prepared data, and created visualizations for this study. P.I.M. and A.T.W. drafted the manuscript. All authors reviewed, edited, and approved the final version.

