## Supplemental figures for "Inactivated Japanese encephalitis virus vaccination imprints fusion loop–biased antibody responses that are attenuated by repeated live-attenuated dengue vaccination"

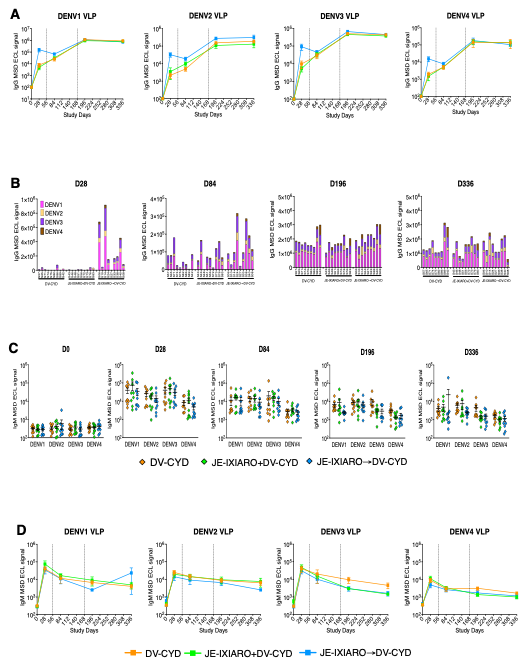

**Fig. S1. Longitudinal DENV IgG and IgM cross-reactive antibody responses following CYD-TDV vaccination across different participant groups (n=27)**. **(A)** DENV1-4-reactive IgG response kinetics over time. **(B)** Breadth of DENV1-4-reactive IgG binding over time. **(C, D)** Magnitude and response kinetics of DENV1-4-reactive IgM antibodies over time. Vertical dashed lines indicate vaccine dose timings (day 0, Dose 1; day 56, Dose 2; day 168, Dose 3). Data were analyzed with mixed-effects models. P values were obtained via the Kruskal-Wallis test with Dunn’s correction and are shown as means ± SEM. Significance was set at p≤0.05. The groups included JE-naïve (DV-CYD, orange, n=9), simultaneous administration (JE-IXIARO+DV-CYD, green, n=9), and JE IXIARO-priming (JE IXIARO→DV-CYD, blue, n=9). Abbreviations: VLP: Virus-like particle, Ig: Immunoglobulin. D28, day 28; D84, day 84; D196, day 196; D336, day 336; DV or DENV, dengue virus; Ig, immunoglobulin; FLE, fusion loop epitope; MSD, meso scale discovery; ECL, electrochemiluminescence; E, envelope protein; EDIII, E domain III; JE, Japanese encephalitis virus; CYD-TDV, chimeric yellow fever virus-derived tetravalent dengue virus vaccine.

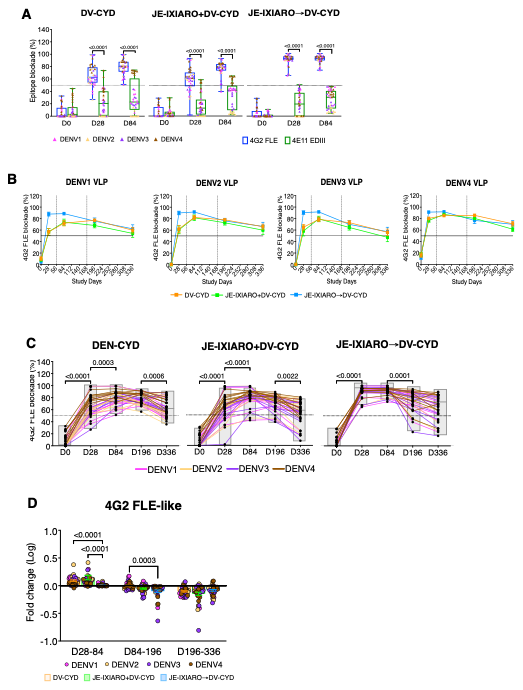

**Fig. S2. Responses of 4G2 FLE- and 4E11 EDIII-like antibodies after CYD-TDV vaccination across different participant groups (n=27)**. **(A) C**omparison of the response magnitude of 4G2 FLE and 4E11 EDII-like antibodies from day 0 to day 84. **(B, C)** Kinetics of 4G2 FLE-like antibody response over time. **(D)** Changes in 4G2 FLE-like antibody response between days 28 and 84, days 84 and 196, and days 196 and 336. Vertical dashed lines mark vaccine dose timings (day 0, Dose 1; day 56, Dose 2; day 168, Dose 3). Data analyzed using mixed-effects models. P values from the Kruskal-Wallis test with Dunn’s correction are shown as means ± SEM. Significance threshold set at p≤0.05. The groups include JE-naïve (DV-CYD, orange, n=9), simultaneous administration (JE-IXIARO+DV-CYD, green, n=9), and JE IXIARO-priming (JE IXIARO-DV-CYD, blue, n=9). All MSD’s experiments were performed as single replicates, utilizing MSD's high throughput, sensitivity, and broad dynamic range. Abbreviations: VLP: virus-like particle, Ig: Immunoglobulin. D28, day 28; D84, day 84; D196, day 196; D336, DAY 336; DV or DENV, dengue virus; Ig, immunoglobulin; FLE, fusion loop epitope; MSD, meso scale discovery; ECL, electrochemiluminescence; E, envelope protein; EDIII, E domain III; JE, Japanese encephalitis virus; CYD-TDV, chimeric yellow fever virus-derived tetravalent dengue virus vaccine.

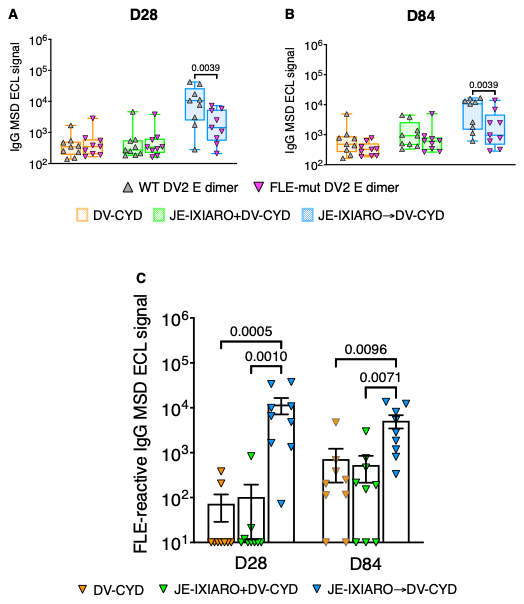

**Fig. S3. IgG response to fusion loop epitope (FLE)-dependent after CYD-TDV vaccination across different participant groups (n=27)**. **(A, B) C**omparison of IgG antibody levels recognizing wild-type E dimers versus mutant E dimers with a G106 amino acid change at days 28 and 84. The FLE-mut E dimers have a specific mutation that reduces the accessibility of the fusion-loop epitope while maintaining the overall E-dimer structure**. (C)** Response magnitude of FLE-reactive IgG at days 24 and 84, calculated by multiplying total IgG binding by the % inhibition of 4G2 FLE-like antibody. P values from Kruskal-Wallis with Dunn’s correction are shown as means ± SEM. Significance is set at p≤0.05. Groups include JE-naïve (DV-CYD, orange, n=9), simultaneous administration (JE-IXIARO+DV-CYD, green, n=9), and JE IXIARO-priming (JE IXIARO-DV-CYD, blue, n=9). All MSD’s experiments were performed as single replicates, utilizing MSD's high throughput, sensitivity, and broad dynamic range. Abbreviations: D28, day 28; D84, day 84; DV or DENV, dengue virus; VLP, virus-like particle; Ig, immunoglobulin; FLE, fusion loop epitope. Mut, mutant; MSD, meso scale discovery; ECL, electrochemiluminescence; WT, wild type; E, envelope protein; JE, Japanese encephalitis virus; CYD-TDV, chimeric yellow fever virus-derived tetravalent dengue virus vaccine.

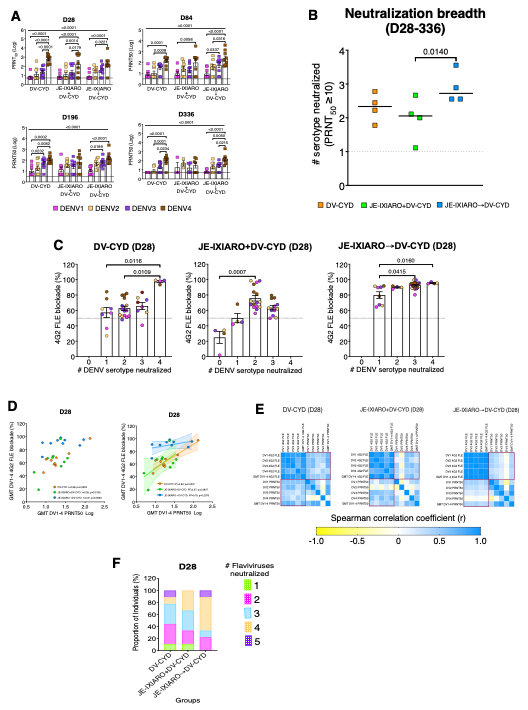

**Fig. S4. Correlation between 4G2 FLE-like antibody levels and neutralization potency, breadth after CYD-TDV vaccination across different groups (n=27)**. **(A)** Neutralization titers against DENV were similar across all three groups and serotypes. **(B, C)** Comparison of the number of DENV1-4 serotypes neutralized (with neutralization breadth defined as PRNT50>10) in relation to 4G2 FLE-like levels and vaccine history/timing over time. **(D, E)** Spearman correlation and regression analysis between the geometric mean of DENV1-4 4G2 FLE-like antibody and DENV neutralization breadth at day 28. **(F)** Neutralization breadth in proportion of individuals across groups. P values above the top bracket are from Kruskal-Wallis tests with Dunn’s correction for multiple comparisons, shown as means ± SEM. A p-value of ≤0.05 indicates significance. Data are presented as geometric mean titers, with individual data points shown. Groups include JE-naïve (DV-CYD, orange, n=9), simultaneous administration (JE-IXIARO+DV-CYD, green, n=9), and JE IXIARO-priming (JE IXIARO-DV-CYD, blue, n=9). All MSD’s experiments were performed as single replicates using MSD's high throughput, sensitivity, and broad dynamic range, whereas all ADE’s experiments were performed in technical duplicates. Abbreviations: D28, day 28; Ig, immunoglobulin; GMT, geometric mean titer; JE, Japanese encephalitis virus; DV or DENV, dengue virus; CYD-TDV, chimeric yellow fever virus-derived tetravalent dengue vaccine; FLE, fusion loop epitope; MSD, meso scale discovery; VLP, virus-like particle; Ig, immunoglobulin; E, envelope protein; PRNT, plaque reduction neutralization test.

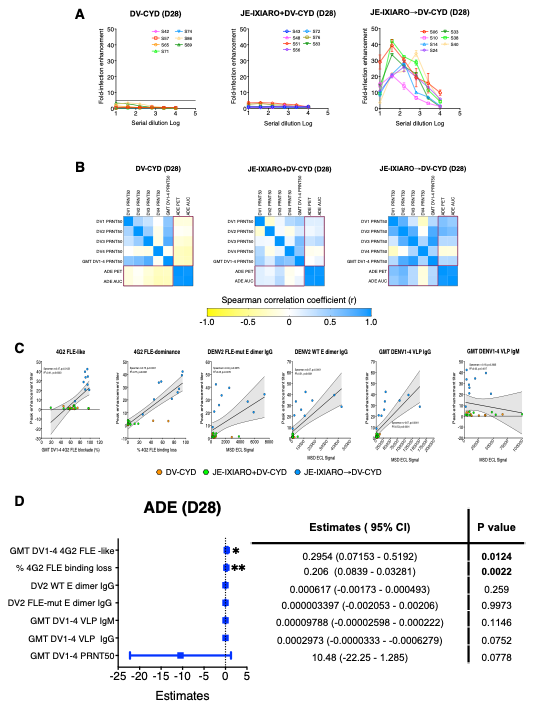

**Fig. S5. Fusion loop–biased imprinting boosts antibody-dependent enhancement (ADE)** a**fter CYD-TDV vaccination in various groups (n=27)**. **(A)** ADE curves using FcγRIIa-expressing K562 cells at 28 days post-vaccination. Serum was mixed with DENV2 RVPs before infection**. (B)** Heatmap displaying correlations between DENV1-4 neutralizing titers and ADE, shown as area under the enhancement curve (AUC) and peak enhancement titer (PET). **(C, D)** Both univariate (scatterplot) and multivariate (forest plot) analyses assess the link between 4G2 FLE-like antibody levels (and percentage of 4G2 FLE loss) and ADE activity, either alone or with other predictors at day 28. Coefficients, p-values (top right of panel C, left of panel D), and linear regression lines with 95% confidence intervals (dotted gray line and shading) are included. A p-value ≤0.05 is significant. Groups include JE-naïve (DV-CYD, orange, n=9), simultaneous administration (JE-IXIARO+DV-CYD, green, n=9), and JE IXIARO-priming (JE IXIARO-DV-CYD, blue, n=9). All MSD’s experiments were performed as single replicates using MSD's high throughput, sensitivity, and broad dynamic range, whereas all ADE’s experiments were performed in technical duplicates. Abbreviations: D28, day 28; Ig, immunoglobulin; GMT, geometric mean titer; JE, Japanese encephalitis virus; DV or DENV, dengue virus; CYD-TDV, chimeric yellow fever virus-derived tetravalent dengue vaccine; FLE, fusion loop epitope; MSD, meso scale discovery; VLP, virus-like particle; Ig, immunoglobulin; E, envelope protein; PRNT, plaque reduction neutralization test.

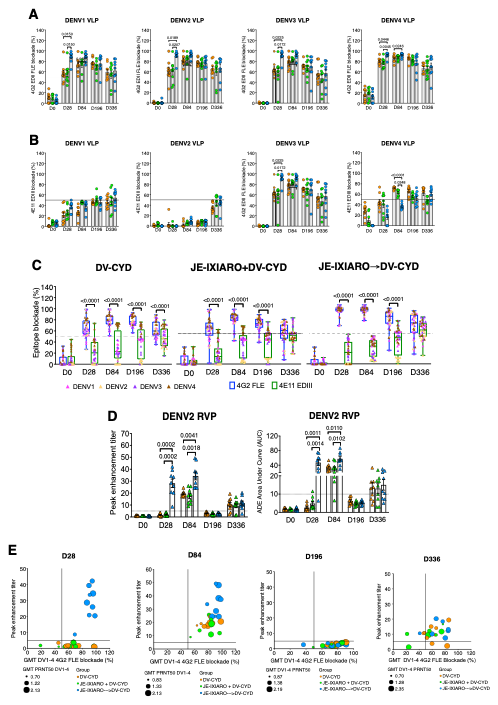

**Fig. S6: Longitudinal analysis of the relationship between 4GE FLE-antibody response, neutralization potency, ADE activity, and vaccine history and timing. (A, B)** Levels of 4G2 FLE-like and 4E11 EDIII-like antibodies following CYD-TDV vaccination across participants over time. **(C)** Comparison of the response magnitude for 4G2 FLE-like and 4E11 EDIII-like antibodies over time. **(D)** ADE activity was quantified as peak enhancement titer (PET) and area under the enhancement curve (AUC) over time. **(E)** Correlation between fusion loop dependence, dengue virus neutralization potency, and FcγRIIa-mediated ADE activity over time. Data analyzed with mixed-effects models. P values are from Kruskal-Wallis tests with Dunn’s correction, shown as means ± SEM. Significance set at p≤0.05. Groups include JE-naïve (DV-CYD, orange, n=9), simultaneous administration (JE-IXIARO+DV-CYD, green, n=9), and JE IXIARO-priming (JE IXIARO-DV-CYD, blue, n=9). All MSD’s experiments were performed as single replicates using MSD's high throughput, sensitivity, and broad dynamic range, whereas all neutralization and ADE’s experiments were performed in technical duplicates. Abbreviations: DV or DENV, dengue virus; JE, Japanese encephalitis virus; VLP, virus-like particle; RVP, reporter virus-particle; FLE, fusion loop epitope; EDIII, E domain III; CYD-TDV, chimeric yellow fever virus-derived tetravalent dengue virus vaccine; ADE, antibody-dependent enhancement; GMT, geometric mean titer; MSD, meso scale discovery; E, envelope protein; ADE, antibody-dependent enhancement; PRNT, plaque reduction neutralization test; SEM, standard error.

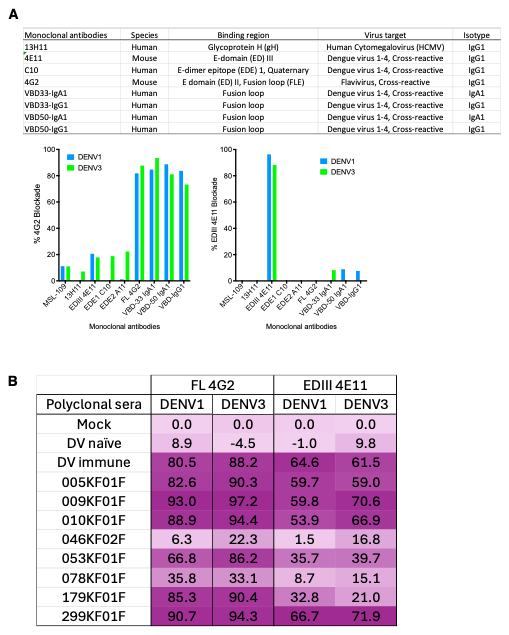

**Fig. S7. Multiplex MSD ECL-based blockade of binding assay evaluation.** **(A and B)** Bar graphs showing percent blockade (Ab) by monoclonal antibody (mAb; A) and polyclonal serum IgG (B) of 4G2 FLE and 4E11 EDIII mAbs binding to DENV1 and DENV3 VLPs. 4G2 FLE-like and 4E11 EDIII-like Ab levels were defined as percent blockade ≥ 50% at 10 µg/ml for mAbs or at 1:10 dilution for polyclonal sera. Mock, control without mAb or sera; 4E11 is a mouse-derived DENV cross-reactive mAb targeting the E domain (ED) III; 4G2 is a mouse-derived DENV cross-reactive mAb targeting the EDII fusion loop epitope (FLE). Abbreviation: DV or DENV, dengue virus; mAb, monoclonal antibody; VLP, virus-like particle; MSD,, meso scale discovery; E, envelope protein; EDIII, E domain III; FLE, fusion loop epitope; Ig, immunoglobulin.
